# Analysis of 21 appendices from children with multisystem inflammatory syndrome compared to specimens of acute appendicitis – a new insight into MIS-C pathology

**DOI:** 10.1101/2022.11.02.22281838

**Authors:** Magdalena Okarska-Napierała, Weronika Woźniak, Joanna Mańdziuk, Kamila Maria Ludwikowska, Wojciech Feleszko, Jakub Grzybowski, Mariusz Panczyk, Elżbieta Berdej-Szczot, Janusz Zaryczański, Barbara Górnicka, Leszek Szenborn, Ernest Kuchar

## Abstract

**Background & Aims:** Multisystem inflammatory syndrome in children (MIS-C) is a rare, but severe complication of coronavirus disease 2019, commonly involving the gastrointestinal tract. Some children with MIS-C undergo appendectomy before the final diagnosis. There are several hypotheses explaining pathomechanism of MIS-C, with the central role of the viral antigen persistence in the gut, associated with lymphocyte exhaustion and immune system dysregulation. We aimed to examine appendectomy specimens obtained from MIS-C patients and analyze the pathological features of the disease and the presence of severe acute respiratory syndrome coronavirus 2 (SARS-CoV-2) antigens in the appendix.

**Methods:** In this cross-sectional study we included 21 children with MIS-C who underwent appendectomy before the final diagnosis. MIS-C patients were recruited from the Polish national registry of inflammatory syndromes in children. The control group included 21 sex- and age-matched children with acute appendicitis (AA) unrelated to SARS-CoV-2 infection. Histological evaluation involved hematoxylin and eosin staining and immunohistochemical identification of lymphocyte subpopulations, programmed cell death protein 1, and SARS-CoV-2 nucleocapsid antigen.

**Results:** Appendices of MIS-C patients lacked neutrophilic infiltrate of muscularis propria typical for AA (14 vs 95%, p<0.001). The proportion of CD20^+^ to CD5^+^ cells was higher in patients with MIS-C (p 0.04), as well as the proportion of CD4^+^ to CD8^+^ (p <0.001). We found no proof of SARS-CoV-2 antigen presence, nor lymphocyte exhaustion, in the appendices of MIS-C patients.

**Conclusions:** Our findings describe pathomorphological features of the appendix in MIS-C and argue against the central role of SARS-CoV-2 persistence in the gut and concomitant lymphocyte exhaustion as the major triggers of MIS-C.

## 1. Introduction

Multisystem inflammatory syndrome in children (MIS-C) is a rare, but severe complication of either symptomatic or asymptomatic infection with the new severe acute respiratory syndrome coronavirus 2 (SARS-CoV-2).^1–5^ MIS-C develops approximately four weeks after coronavirus disease 2019 (COVID-19) in 2-3 per 10.000 infected children.^4,6–8^ MIS-C presents with fever, lethargy, multiorgan damage of varying severity, and laboratory markers of hyperinflammation. A substantial proportion of MIS-C patients necessitate pediatric intensive care unit (PICU) hospitalization, mostly due to cardiovascular complications, with hypotension and myocardial dysfunction.^1–4,9^ Gastrointestinal involvement (nausea, vomiting, diarrhea, and abdominal pain) is particularly common during MIS-C.^2,3,5,9^ A combination of severe abdominal pain and peritoneal signs sometimes leads to surgical interventions for the ‘acute abdomen’ before the final diagnosis is made.^1,5,10–12^

MIS-C is an effect of immune dysregulation, involving activation of macrophages, dendritic cells and neutrophils, profound peripheral lymphopenia and a ‘cytokine storm’, triggered by SARS-CoV-2. However, the exact pathomechanism of the disease is unknown. Prolonged fecal shedding of SARS-CoV-2 antigens in children^13^ and pronounced abdominal symptoms and signs during MIS-C suggest possible role of the intestine in MIS-C pathogenesis. Increased intestinal permeability which could lead to translocation of the viral particles from the gut into the circulation is one of the proposed hypotheses.^14,15^ Another theory is persistent exposure of the immune system to SARS-CoV-2 antigens leading to lymphocyte exhaustion.^16–18^ The histopathologic picture of MIS-C is largely unknown. There are only single case reports or small case series presenting histologic findings of the selected tissues from children with this new entity.^10,11,19–21^

We aimed to examine appendectomy specimens obtained during MIS-C and analyze the pathomorphological features of the disease and the presence of SARS-CoV-2 antigens in the affected tissue.

## 2. Material and methods

### 2.1. Patients’ recruitment

In this retrospective observational study, we have analyzed the appendix tissue specimens obtained during appendectomies. The study was approved by the Bioethical Committee of Medical University of Warsaw (Approval No: KB/135/2021).

The study group (MIS-C group) involved children who underwent appendectomy during MIS-C. We searched the national registry of inflammatory diseases in children (the MultiOrgan Inflammatory Syndromes Covid Related registry, MOIS-CoR)^5^ for patients under the age of 18 years old, with MIS-C diagnosed according to the World Health Organization (WHO) definition, who had undergone appendectomy after the onset of MIS-C symptoms and signs, but before immunomodulatory therapy was introduced. Patients’ demographic, clinical and laboratory data were obtained from the MOIS-CoR registry. Children with MIS-C included in the study were operated between July 2020 and February 2022.

The control group involved sex- and age-matched children with acute appendicitis (AA), who underwent appendectomy in the Pediatric Teaching Clinical Hospital of the Medical University of Warsaw and did not have a history of immunomodulatory treatment or immunodeficiencies. Patients with AA and positive polymerase chain reaction (PCR) for SARS-CoV-2 in the nasopharynx were excluded from the study. Patients’ demographic, clinical and laboratory data were obtained from the hospital electronic medical records. Children with AA included in the study were operated between July 2020 and March 2022.

### 2.2. Study definitions

MIS-C was diagnosed based on WHO case definition,^22^ which included all of the following:

- Age <18 years old
- Fever ≥3 days
- Elevated inflammatory markers, at least one of the following: erythrocyte sedimentation rate (ESR), C-reactive protein (CRP), or procalcitonin (PCT)
- No other identified microbial cause of inflammation
- Evidence of COVID-19 (PCR, antigen test or serology positive, or likely contact with COVID-19 case)
- At least two of the following:
  - Rash or bilateral non-purulent conjunctivitis or mucocutaneous inflammation signs (oral, hands, or feet)
  - Hypotension
  - Features of myocardial dysfunction (decreased left ventricular ejection fraction [LVEF], pericarditis, or coronary abnormalities – based on echocardiographic findings, or elevated troponin/ B-type natriuretic peptide [BNP] or N-terminal prohormone of BNP [NT-proBNP])
  - Evidence of coagulopathy (an increase of at least one of the following: international normalized ratio [INR], activated partial thromboplastin time [APTT], D-dimer)
  - Acute gastrointestinal symptoms (diarrhea, vomiting, or abdominal pain).

Laboratory results on admission to the hospital and at their respective peaks (if available) were included in the analysis. Elevated ESR was defined as >40 mm/hour, elevated CRP as >30 mg/L, and elevated PCT as >0.5 ng/mL. Elevated INR was defined as >1.1, elevated APTT as >40 s, and elevated D-dimer as >500 ug/L. Elevated troponin I or T was defined as >50 ng/L and elevated BNP or NT-proBNP as >150 pg/mL.

Hypotension was defined as the minimum systolic blood pressure (sBP) of below (70 + 2× [age in years]) mmHg for children ≤10 years old and <90 mmHg for children >10 years old.^23^ Decreased LVEF was defined as the minimum LVEF below 55%.

Overweight was defined as body mass index (BMI) over 85th percentile and obesity - as BMI over 95th percentile, according to the Centers for Disease Control and Prevention (CDC) percentile charts.^24^

### 2.3. Tissue preparation and evaluation

After obtaining written informed consent from children’s carers, we have collected appendix specimens in the form of paraffin blocks from the hospitals where children were operated. The blocks were cut into 3-5 μm thick sections and were stained in an automatic tissue processor with H&E. Subsequent, corresponding sections were stained with the following antibodies: anti-CD5 (clone 4C7, DAKO/Agilent, USA), anti-CD20cy (clone L26, DAKO/Agilent, USA), anti-CD4 (clone 4B12, DAKO/Agilent, USA), anti-CD8 (clone C8/144B, DAKO/Agilent, USA), anti-PD-1 (clone NAT105, Ventana/ROCHE, USA) and anti-SARS-CoV-2 (clone BSB-134, Bio SB, USA). All the stainings were performed in accordance with the antibodies producers’ recommendations and EnVision FLEX, High pH (Link) (DAKO/Agilent, USA) and Autostainer Link 48 (DAKO/Agilent, USA) were used to visualize the effects.

Analysis of both the hematoxylin and eosin (H&E) stainings and the immunohistochemical results was performed by a single pathologist using a light microscope.

The neutrophilic infiltration - a histological marker of AA^25^ - was evaluated as either present or absent in three layers of the appendix wall: (i) mucosa/submucosa, (ii) muscularis propria and (iii) serosa.

To compare the abundance of lymphocytes B (CD20^+^ cells) and lymphocytes T (CD5^+^ cells) within the wall of each appendix, corresponding stainings were assessed with the use of low magnification (objective ×4). Assuming that both of these cell populations constitute 100% of examined cells on both slides together, an appropriate percentage was assigned to each population. The same was done to assess the two subpopulations (CD4^+^ and CD8^+^) of T lymphocytes.

Programmed cell death 1 protein (PD-1) was used as a marker of lymphocyte activation and exhaustion.^18,26^ Cells expressing this protein were sought within lymphoid tissue of each appendix. As this protein is also highly expressed in normal T follicular helper cells (Tfh), a truly positive reaction was supposed to be present beyond germinal centers. The presence of Tfh was treated as a positive internal control.

The presence of SARS-CoV-2 nucleocapsid protein was sought in the epithelia, mesothelia, endothelia and myocytes. Since weak staining of this antibody is commonly seen in some plasma cells (regardless of real presence of the antigen) these cells were not counted as a positive result, but only as a positive internal control. Moreover, because anti-SARS-CoV-2 antibody is assigned to research purposes only and is not used in routine diagnostic procedures, its pattern of staining was tested on control slides provided by the producer (Bio SB, USA).

### 2.4. Statistical analysis

Data were presented using descriptive statistics parameters according to the type of variable: median (Mdn), quartiles 1 and 3 (Q1-Q3), count (n), and frequency (%). Differences between groups (MIS-C vs AA) and correlations between clinical variables were assessed using statistical inference (null hypothesis testing). The non-parametric data analysis was chosen due to the small sample size.^27^ The two-sided Fisher’s exact test (for dichotomous variables) and the Mann-Whitney-Wilcoxon test with continuity correction (for ordinal or continuous variables) were used. The correlation analysis was performed by determining the Spearman’s rank correlation coefficient. For all analysis, a priori statistical significance level was set to 0.05. Data were analyzed using STATISTICA version 13.3 (Tibco Software Inc., Palo Alto, California, USA) under the license of the Medical University of Warsaw.

## 3. Results

### 3.1. Characteristics of the study and control groups

The analysis involved 21 MIS-C patients and 21 AA patients. Initially we have recruited 22 children with MIS-C and 22 children with AA. One boy was at first hospitalized with fever, peritoneal signs, conjunctivitis, low lymphocyte count (0.8 ×10^3^/µL), high inflammatory markers and PCR for SARS-CoV-2 (cycle threshold of 19.1 for gene E and 20.8 for N2). He received ceftriaxone and metronidazole and underwent appendectomy five days after symptoms onset. The fever subsided four days after surgery. Two weeks later the boy developed sudden deterioration with high fever, hyperinflammation and hypotension, and was treated for MIS-C. We have examined his appendix specimen, but due to the unclear character of his disease at the time of surgery, the boy was excluded from the analysis and so was the matching patient from the AA group.

Demographic and clinical features of both groups, as well as vital signs and laboratory results on admission to the hospital, are presented in Table 1. Children with MIS-C did not differ from children with AA in terms of abdominal symptoms and signs, but mucocutaneous abnormalities were absent in patients with AA and fever was more common in MIS-C. On admission, children with MIS-C had lower median sBP (104 vs 108 mmHg, p 0.04) and 23% of them were hypotensive, whereas no children with AA had decreased blood pressure – however, the latter difference did not reach statistical significance. There were significant differences between the groups in some laboratory results on admission to the hospital: children with MIS-C had lower white blood cell, lymphocyte and platelet counts, and higher CRP. Moreover, patients with MIS-C initially presented with low albumin concentration, high PCT, and high BNP/NT-proBNP – those parameters were not available in the AA group.

**Table 1.** Demographic and clinical features, and selected parameters on admission to the hospital of patients with multisystem inflammatory syndrome in children (MIS-C) and acute appendicitis (AA).

| Clinical and laboratory characteristics<br>n/N (%) or median (Q1-Q3) |  | MIS-C<br>(n=21) | AA (n=21) | p* |
| --- | --- | --- | --- | --- |
| Demographics | Age [years] | 8 (6.1-10.8) | 7.9 (6.4-10.9) | NS |
|  | Sex [boys] | 11/21 (52) | 12/21 (57) | NS |
|  | BMI [kg/m<sup>2</sup>]** | 16.5 (14.2-18.9) | 15.3 (14-16.7) | NS |
|  | Comorbidities*** | 1/19 (5) | 2/21 (10) | NS |
| Clinical presentation | **Fever** | **21/21 (100)** | **8/21 (38)** | **<0.001** |
|  | Abdominal pain | 21/21 (100) | 21/21 (100) | NS |
|  | Diarrhea | 7/21 (33) | 9/21 (43) | NS |
|  | Nausea/vomiting | 16/21 (76) | 15/21 (71) | NS |
|  | **Rash** | **19/21 (95)** | **0/21 (0)** | **<0.001** |
|  | **Conjunctivitis** | **14/21 (66)** | **0/21 (0)** | **<0.001** |
|  | **Oral inflammation** | **12/20 (60)** | **0/21 (0)** | **<0.001** |
|  | **Distal extremities changes** | **17/21 (81)** | **0/21 (0)** | **<0.001** |
|  | **Cervical lymphadenopathy** | **5/21 (24)** | **0/21 (0)** | **0.048** |
| Vitals on admission | HR [/min] | 126 (105-133) | 116 (93-131) | NS |
|  | **sBP [mmHg]** | **104 (99-106)** | **108 (103-119)** | **0.038** |
|  | Hypotension | 3/14 (23) | 0/13 (0) | NS |
| Laboratory results on admission | **WBC [$\times 10^3/\mu\text{L}$]** | **8 (4.7-13.2)** | **14.9 (9.1-18.1)** | **0.02** |
| | Neutrophils [$\times 10^3/\mu\text{L}$] | 7.6 (3.9-10.2) | 11.1 (7.1-15.2) | NS |
|  | **Lymphocytes [$\times 10^3/\mu\text{L}$]** | **0.8 (0.4-1.2)** | **1.6 (1.2-2)** | **0.002** |
|  | **Hemoglobin [g/dL]** | **12 (11.4-12.5)** | **13.1 (12.6-13.5)** | **0.002** |
|  | **Platelets [$\times 10^3/\mu\text{L}$]** | **160 (132.3-198.3)** | **308 (250-370)** | **<0.001** |
|  | Sodium [mmol/L] | 134 (134-136.5) | 138 (136.5-138.5) | NS |
|  | **CRP [mg/L]** | **147.5 (69.5-205.4)** | **52 (29.5-71.2)** | **<0.001** |
|  | PCT [ng/mL] | 3.5 (1.4-9.5) | - | - |
|  | Albumin [g/dL] | 2.8 (2.2-3.6) | - | - |
|  | BNP/NT-proBNP [pg/mL] | 4426 (850-13597) | - | - |
| SARS-CoV-2 tests | SARS-CoV-2 PCR positive | 3/18 (17) | 0/21 (0) | - |
|  | SARS-CoV-2 serology positive | 20/20 (100) | - | - |

AA, acute appendicitis; BMI, body mass index; BNP, B-type natriuretic peptide; CRP, C-reactive protein; HR, heart rate; MIS-C, multisystem inflammatory syndrome in children; NS, not significant; NT-proBNP, N-terminal prohormone of B-type natriuretic peptide; PCR, polymerase chain reaction; PCT, procalcitonin; Q1, first quartile; Q3, third quartile; SARS-CoV-2, severe acute respiratory syndrome coronavirus 2; sBP, systolic blood pressure; WBC, white blood cells.
\*Categorical variables analyzed using Fisher's exact test, continuous variables by Mann-Whitney-U test with continuity correction.
\*\*One case of obesity and three cases of overweight in the MIS-C group; one case of obesity and one case of overweight in the AA group.
\*\*\*One case of asthma in the MIS-C group; two cases of asthma in the AA group.

All children underwent appendectomy. Among patients with AA, all but one had a laparoscopy, whereas in the MIS-C group, nine children (43%) had a laparoscopy and 12 (57%) had a laparotomy. The median time from symptoms onset to hospitalization was three (range 0-5) days in MIS-C patients and one (range 0-3) day in AA patients (p 0.002), whereas the median time from admission to the hospital to the operation was one (range 0-5) day in MIS-C and one (range 0-4) day in AA (p 0.32).

The clinical course of MIS-C, with the most abnormal laboratory and imaging findings, is presented in Table 2. The corresponding laboratory results at their respective peaks from children with AA are included when available. All children from both groups fully recovered.

**Table 2.** Clinical course of multisystem inflammatory syndrome in children (MIS-C) in the study group and selected results of laboratory tests at their respective peaks in the acute appendicitis (AA) group.

| **Clinical course of MIS-C**<br>n/N (%) or median (Q1-Q3) |  | MIS-C<br>(n=21) | AA (n=21) | p* |
| --- | --- | --- | --- | --- |
| Blood pressure and imaging findings at respective peaks | sBP, the lowest [mmHg] | 82 (74-91.3) | - | - |
|  | Hypotension | 11/20 (55) | - | - |
|  | LVEF, the lowest [%] | 58 (50-66) | - | - |
|  | Decreased LVEF | 5/18 (28) | - | - |
| Laboratory results at respective peaks | **Lymphocytes min.**<br>[x10<sup>3</sup>/μL] | **0.7 (0.5-0.9)** | **1.4 (1.2-1.7)** | **<0.001** |
|  | **Hemoglobin min.**<br>[g/dL] | **9.2 (8.7-10.3)** | **12.5 (11.7-13.1)** | **<0.001** |
|  | **Platelets min.** [x10<sup>3</sup>/μL] | **130 (91-169)** | **278 (223-321)** | **<0.001** |
|  | **Sodium min.** [mmol/L] | **134 (132-135)** | **137 (136-138)** | **0.004** |
|  | **CRP max.** [mg/L] | **241.6 (145.7-311)** | **78.5 (47.5-131.7)** | **<0.001** |
|  | PCT max. [ng/mL] | 8 (2.3-13) | - | - |
|  | Ferritin [pg/L] | 421.8 (306.3-852.4) | - | - |
|  | Albumin min. [g/dL] | 2.4 (2.2-2.7) | - | - |
|  | BNP/NT-proBNP max. [pg/mL] | 9479 (1348-17731) | - | - |
|  | **INR max.** | **1.3 (1.3-1.4)** | **1.1 (1.1-1.2)** | **<0.001** |
|  | D-dimer [mg/L] | 5.6 (3.1-11.5) | - | - |
| Treatment | IVIG | 18/21 (86) | - | - |
|  | Glucocorticosteroids | 18/21 (86) | - | - |
|  | PICU | 6/21 (29) | - | - |
|  | Mechanical ventilation | 4/21 (19) | - | - |

AA, acute appendicitis; BNP, B-type natriuretic peptide; CRP, C-reactive protein; INR, international normalized ratio; IVIG, intravenous immunoglobulin; LVEF, left ventricular ejection fraction; MIS-C, multisystem inflammatory syndrome in children; NT-proBNP, N-terminal prohormone of B-type natriuretic peptide; PCT, procalcitonin; PICU, pediatric intensive care unit; Q1, first quartile; Q3, third quartile; sBP, systolic blood pressure.
\* Continuous variables analyzed by Mann-Whitney-U test with continuity correction.

### 3.2. Appendix samples evaluation

#### 3.2.1. Hematoxylin and eosin (H&E) staining

Upon morphologic assessment under H&E staining appendices from MIS-C patients were markedly less swollen and did not present with inflammatory infiltration compared to specimens from AA patients. These differences are illustrated in Figure 1.

**Figure 1.**
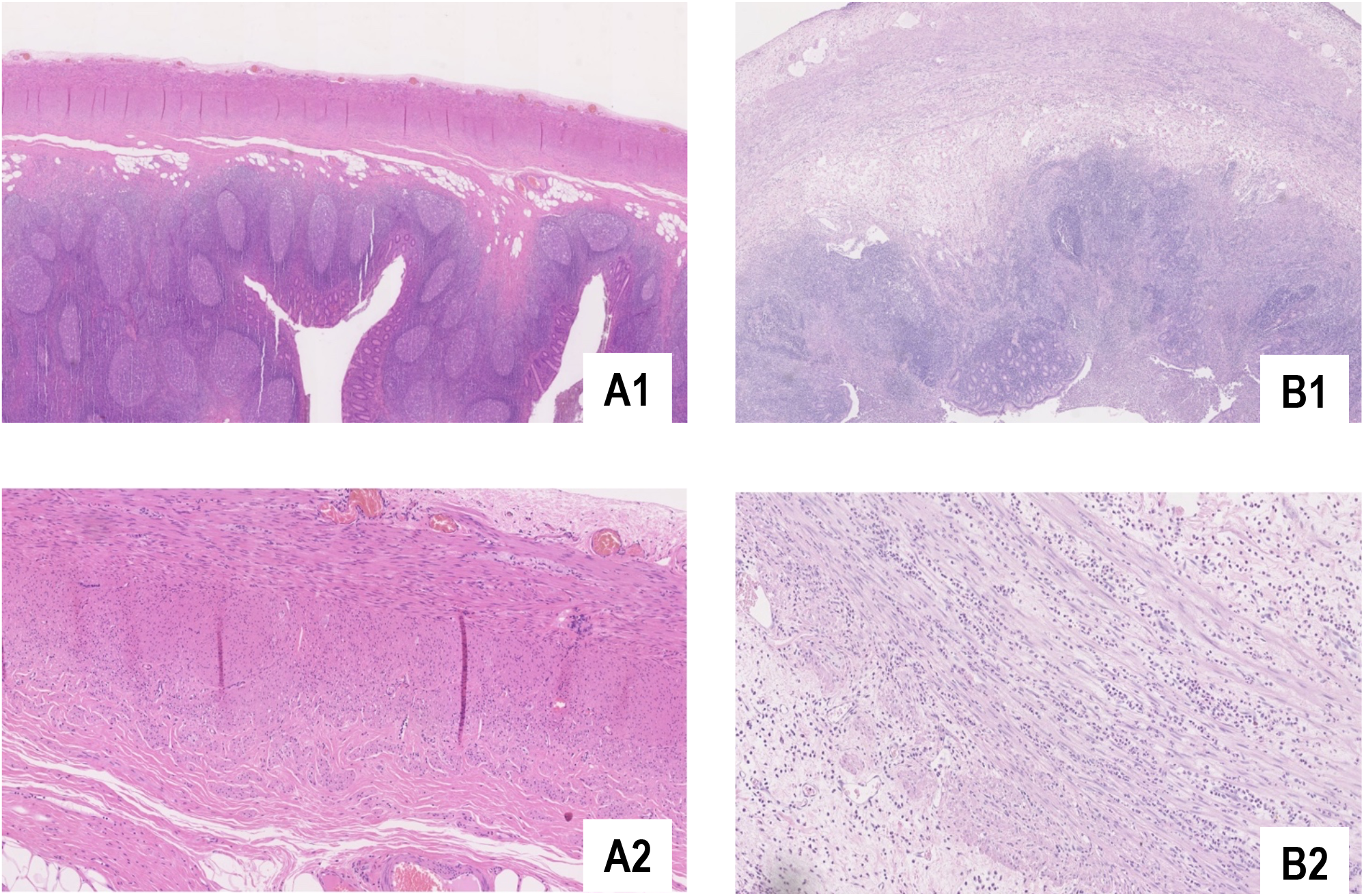
Cross-section of the appendices from a patient with the multisystem inflammatory syndrome in children (MIS-C) (pictures A1 and A2) and a patient with acute appendicitis (AA) (pictures B1 and B2) on H&E stain, under 2x (pictures A1 and B1) and 10x magnification (pictures A2 and B2 - muscularis propria). The appendix wall of the AA patient is edematous, with rich neutrophilic infiltrate, as opposed to the MIS-C patient, whose appendix wall structure looks normal.

Numerical differences in histopathologic findings between studied groups are summarized in Table 3. Neutrophilic infiltrate was significantly more common in AA patients than in MIS-C patients (Figures 1 and 2). Twelve patients with MIS-C (57%) had no neutrophils in the appendix. Neutrophils were absent in muscularis propria of 20/21 patients with MIS-C (95%), as opposed to 3/21 patients with AA (14%, p <0.001). Within the MIS-C group, there was no correlation between neutrophils’ presence in the appendix and laboratory markers of inflammation (lymphocyte count, CRP, PCT, D-dimer, fibrinogen) or clinical markers of the disease severity (decreased sBP, the need for PICU or mechanical ventilation).

**Table 3.** Histopathologic findings in the appendix specimens in patients with multisystem inflammatory syndrome in children (MIS-C) and with acute appendicitis (AA).

| **Histopathological findings in the appendix specimens**<br>n (%) or median (Q1-Q3) |  | MIS-C<br>(n=21) | AA (n=21) | p* |
| --- | --- | --- | --- | --- |
| Neutrophils' presence in the appendix wall layers | **mucosa/submucosa** | **7 (33)** | **19 (90)** | **<0.001** |
|  | **muscularis propria** | **1 (5)** | **18 (86)** | **<0.001** |
|  | **serosa** | **6 (29)** | **18 (86)** | **<0.001** |
| Lymphocyte subpopulations in the appendix specimens | **CD20<sup>+</sup> [%]** | **50 (45-60)** | **45 (45-50)** | **0.041** |
|  | **CD5<sup>+</sup> [%]** | **50 (40-55)** | **55 (50-60)** | **0.041** |
|  | **CD20<sup>+</sup>:CD5<sup>+</sup>** | **1 (0.8-1.5)** | **0.8 (0.7-1)** | **0.041** |
|  | **CD4<sup>+</sup> [%]** | **95 (95-95)** | **90 (80-90)** | **<0.001** |
|  | **CD8<sup>+</sup> [%]** | **5 (5-5)** | **10 (10-20)** | **<0.001** |
|  | **CD4<sup>+</sup>:CD8<sup>+</sup>** | **19 (19-19)** | **9 (4-9)** | **<0.001** |

AA, acute appendicitis; MIS-C, multisystem inflammatory syndrome in children; PD-1, programmed cell death protein 1; Q1, first quartile; Q3, third quartile.
\*Categorical variables analyzed using Fisher's exact test, continuous variables by Mann-Whitney-U test with continuity correction.

**Figure 2.**
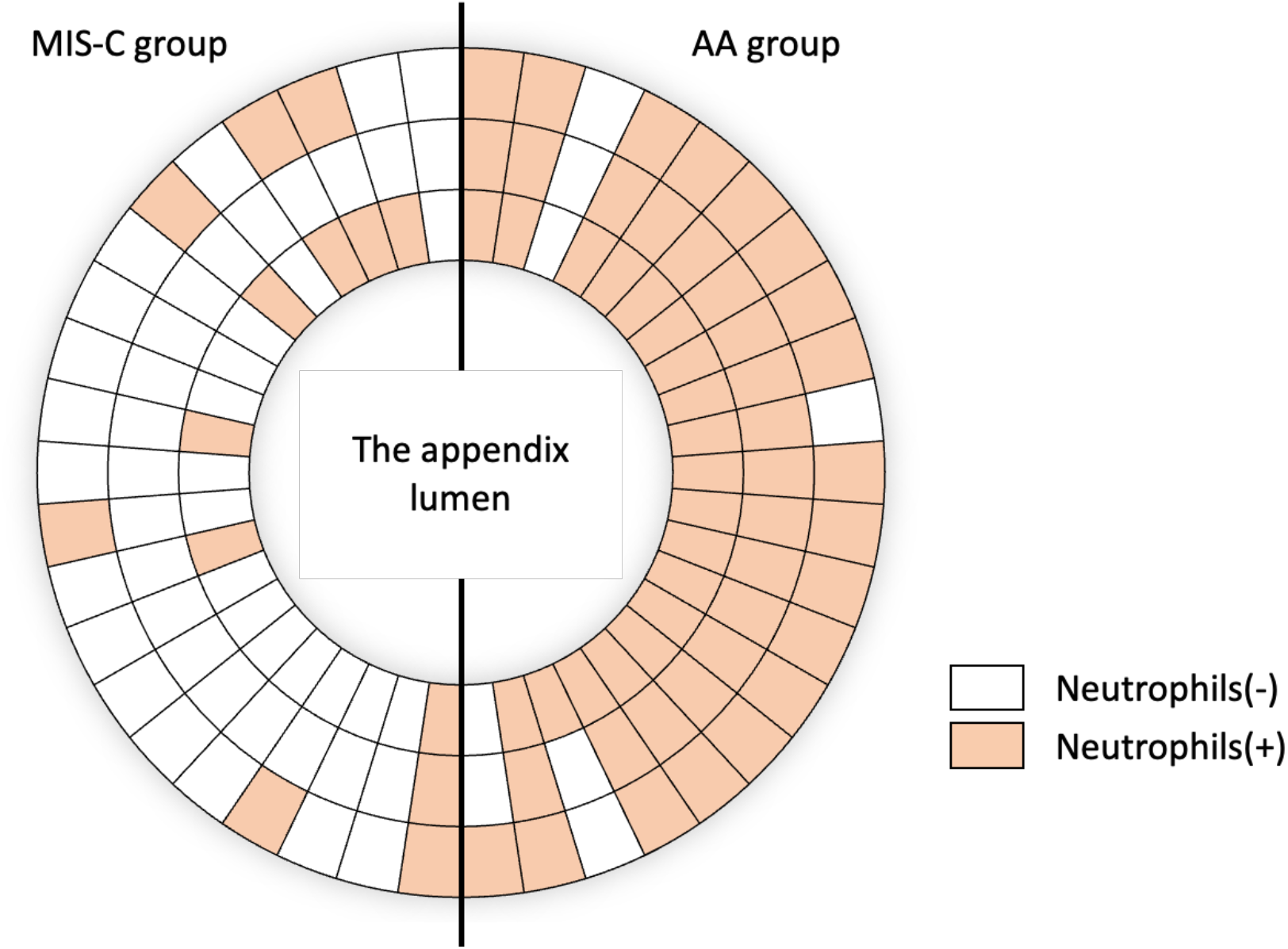
Neutrophils’ presence in the appendix wall layers. Each radius represents a single patient. The inner circle represents the appendix mucosa/submucosa, the intermediate – muscularis propria, and the outer – serosa. AA, acute appendicitis; MIS-C, multisystem inflammatory syndrome in children.

#### 3.2.2. Immunohistochemical staining

On the immunohistochemical assessment of lymphocyte subpopulations, we found significant differences in CD20^+^ to CD5^+^ and CD4^+^ to CD8^+^ proportions between children with MIS-C and AA (Table 3). The proportion of CD20^+^ to CD5^+^ was higher in patients with MIS-C than with AA (p 0.04). This difference is illustrated in Figure 3. The proportion of CD4^+^ to CD8^+^ was higher in MIS-C patients, too (p <0.001), which is illustrated in Figure 3. Within the MIS-C group, there was no correlation between either CD20^+^ to CD5^+^ or CD4^+^ to CD8^+^ ratios in the appendix and laboratory markers of inflammation (lymphocyte count, CRP, PCT, D-dimer, fibrinogen) or clinical markers of the disease severity (decreased sBP, the need for PICU or mechanical ventilation).

**Figure 3.**
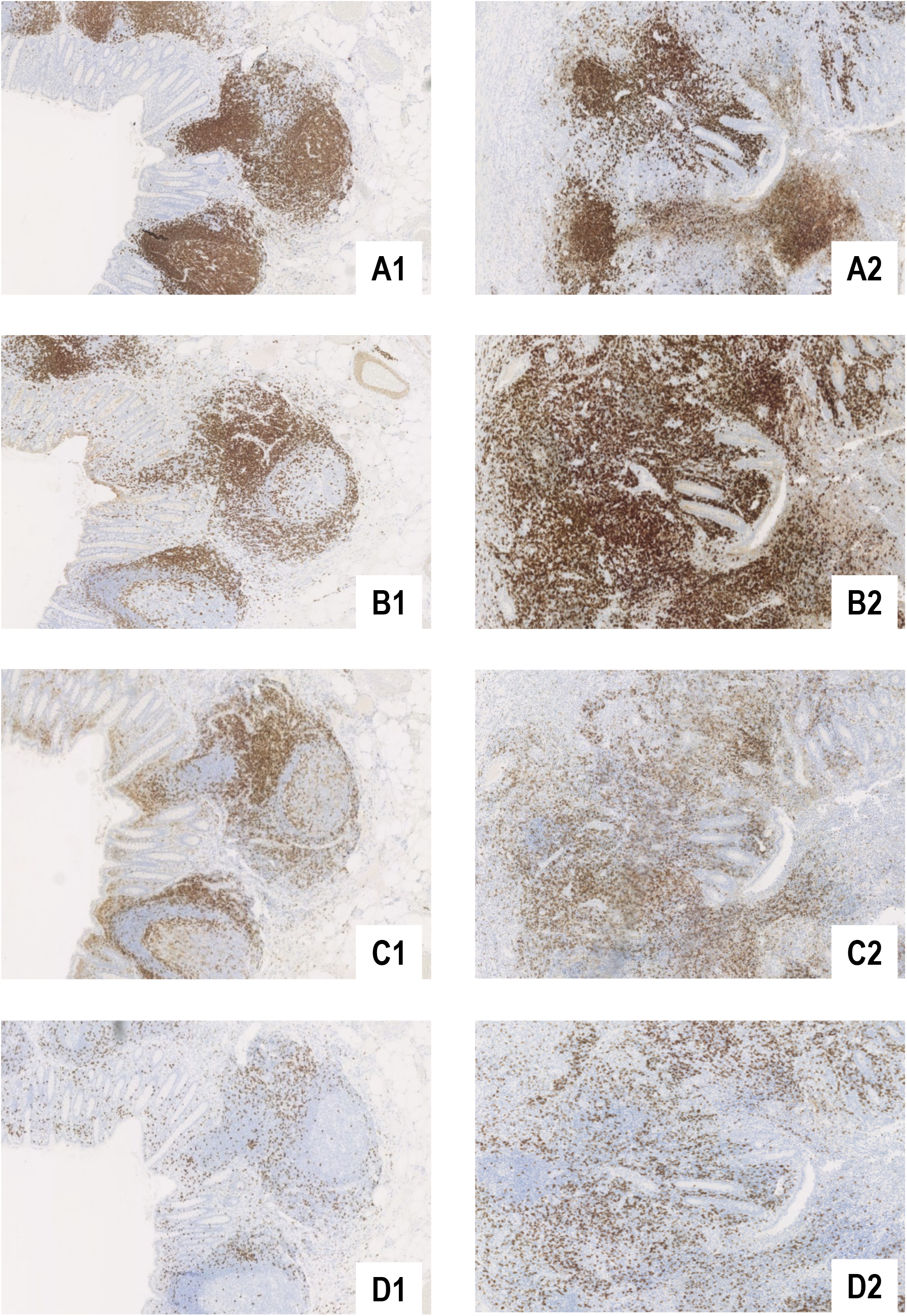
Immunohistochemical identification of lymphocyte subpopulations: CD20^+^ (pictures A1 and A2), CD5^+^ (pictures B1 and B2), CD4^+^ (pictures C1 and C2) and CD8^+^ (pictures D1 and D2) cells in the appendix wall of a patient with the multisystem inflammatory syndrome in children (MIS-C) (pictures A1-D1) and of a patient with acute appendicitis (AA) (pictures A2-D2). CD20^+^ cells (lymphocytes B) predominate in the patient with MIS-C (compare pictures A1 and B1), whereas CD5^+^ cells (lymphocytes T) predominate in the patient with AA (compare pictures A2 and B2). The proportion of lymphocytes T CD4^+^ to CD8^+^ is higher in the patient with MIS-C (compare pictures C1 and D1) than in the patient with AA (compare pictures C2 and D2). Lymphocytes CD8^+^ are scarce in the MIS-C specimen (picture D1).

PD-1 was expressed in Tfh in all but two children in the MIS-C group and all but one in the AA group. No PD1^+^ cells were found beyond germinal centers in both groups. This is illustrated in Figure 4. No SARS-CoV-2 nucleocapsid antigen reactivity was found in the epithelium, mesothelium, endothelium, or myocytes in both groups. This is illustrated in Figure 4.

**Figure 4.**
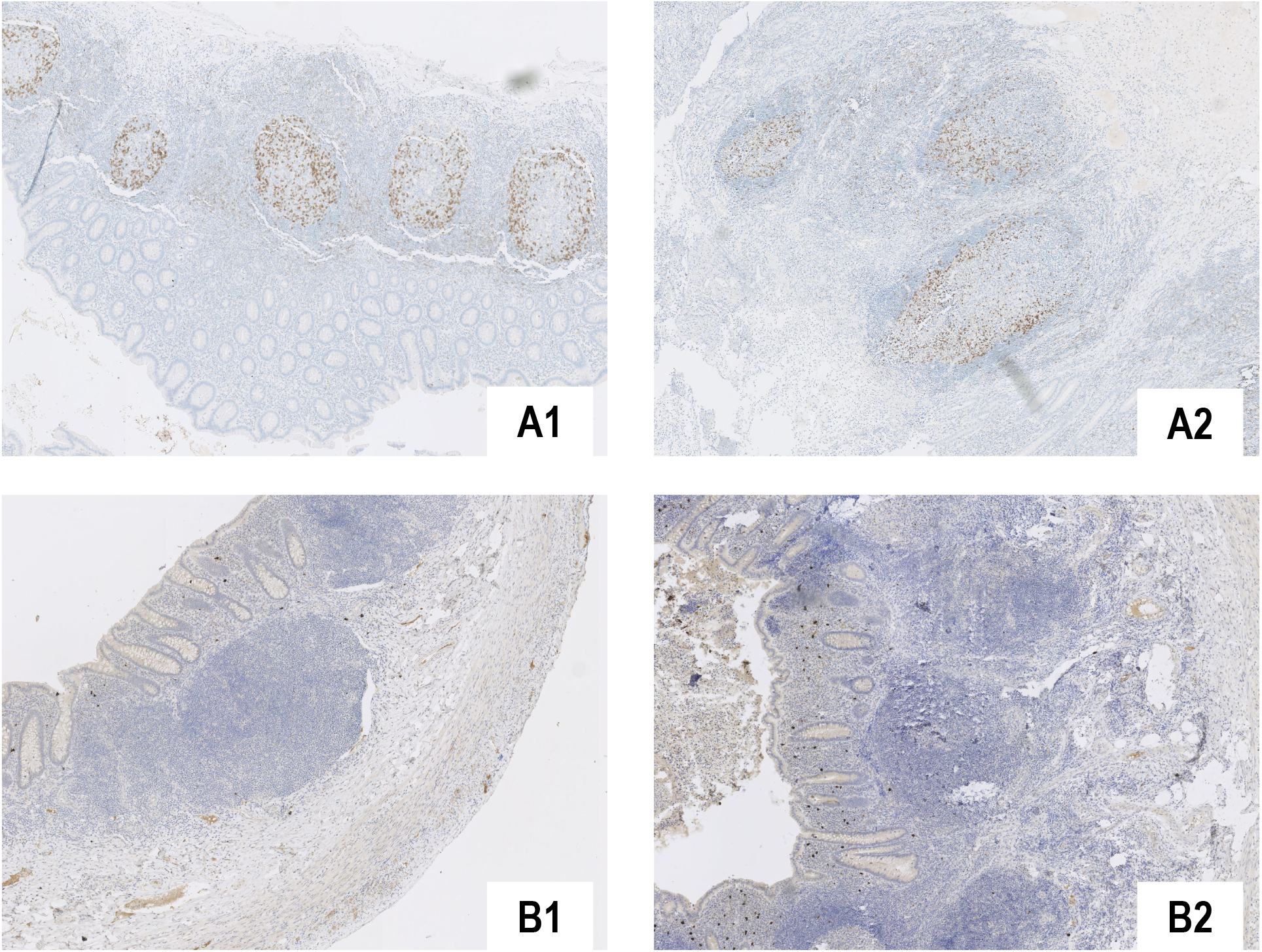
PD-1 staining in the appendix of a patient with the multisystem inflammatory syndrome in children (MIS-C) (picture A1) and a patient with acute appendicitis (AA) (picture A2). PD-1 highlights only T follicular helper cells in the germinal centers and no reaction in other lymphocytes neither in the MIS-C group nor in the AA group, was noted. SARS-CoV-2 staining in the appendix of a patient with multisystem inflammatory syndrome in children (MIS-C) (picture B1) and a patient with acute appendicitis (AA) (picture B2). SARS-CoV-2 was not found in any part of the appendiceal wall. The positive reaction in scattered plasma cells is regarded as non-specific.

## 4. Discussion

Here we describe the largest collection of appendix specimens obtained from children with MIS-C. We found significant clinical, laboratory and pathological differences between MIS-C and AA. The appendices of patients with MIS-C lack edema and neutrophilic infiltrate typical for AA and are characterized by distinct shifts in lymphocyte subpopulations. Moreover, we found no proof of SARS-CoV-2 antigen presence nor lymphocyte exhaustion in the appendices of MIS-C patients, which suggests that the appendix is not likely to be the site of persist antigen expression.

### 4.1. Distinct clinical and laboratory features of MIS-C

It is well documented that 10-20% of children with MIS-C may present with the ‘acute abdomen’.^12,28^ This might be due to peritonitis and mesenteric lymphadenitis, which are commonly reported in patients operated during MIS-C.^28^ Thus, identifying features distinguishing MIS-C from other surgical emergencies (AA being the most common) is an important clinical issue. Our patients with MIS-C were distinguished by some associating signs, such as rash, conjunctivitis, or decreased sBP, and distinct laboratory findings. Moreover, children with MIS-C were admitted to the hospital later since the onset of symptoms compared to patients with AA. Most of these findings are consistent with another study comparing MIS-C to AA, which revealed that longer abdominal pain and fever duration, higher neutrophil count, lower lymphocyte count, and higher CRP were sensitive markers of MIS-C.^29^ Comparing patients with MIS-C and AA, Azılı et al. analyzed laboratory findings at their respective peaks during hospitalization, whereas we found several distinctive features of MIS-C present already on admission to the hospital. These may facilitate early differential diagnosis. Lower sBP, very high inflammatory markers, and a relatively low white blood cell count associated with lymphopenia, hypoalbuminemia and elevated BNP/NT-proBNP should raise the suspicion of MIS-C.

Children from our MIS-C cohort presented a typical combination of hyperinflammation and multisystem injury. However, it is noteworthy that compared to the cohort of 274 children with MIS-C from the Polish MOIS-CoR registry,^5^ our group had a more severe course of the disease, with 29% hospitalized in PICU and 19% necessitating mechanical ventilation (as opposed to 8% and 4%, respectively, in the Polish MIS-C cohort). It can be hypothesized that children with MIS-C associated with the ‘acute abdomen’ represent the more severe spectrum of the disease, as was suggested by Abrams et al., who found that abdominal pain was a risk factor for PICU hospitalization in MIS-C.^30^ However, on the other hand, surgery performed during evolving hyperinflammation could have worsened the course of the disease, which emphasizes the importance of the early differential diagnosis to avoid unnecessary invasive procedures.

Nevertheless, MIS-C may be associated with intestinal inflammation severe enough for a surgical intervention to be necessary. According to a review by Rouva, among 35 children with MIS-C and ‘acute abdomen’ who underwent an operation, 51% of surgeries were proven unnecessary, whereas the rest (49%) revealed true abdominal surgical emergencies.^12^ This must be interpreted with caution because surgical emergencies were not precisely defined nor verified. Nevertheless, there are reports of intestinal perforation during MIS-C,^31–33^ so the diagnosis of MIS-C cannot exclude the need for surgery.

### 4.2. Scarce neutrophilic infiltration in the appendices of MIS-C patients

Neutrophilic infiltration of the muscularis propria is a hallmark of suppurative appendicitis, and such a pathologic presentation is considered explanatory for the patient’s symptoms of the right iliac fossa pain.^34^ Interestingly, despite clinical signs suggestive of AA, most of our patients with MIS-C had no neutrophils in the appendiceal muscularis. Several other authors noticed similar findings in single case reports and small case series.^10,11,35,36^ Correspondingly, neutrophil blood counts tend to be unchanged in MIS-C,^37^ which contrasts with exceptionally high biochemical inflammatory markers. A similar constellation of a relatively low neutrophil count (within normal limits) and high CRP and PCT was present in our patients. The role of neutrophils in MIS-C pathogenesis is unclear. Boribong et al. found that neutrophils isolated from fresh whole blood of patients with MIS-C express signatures of altered metabolism, activation, degranulation, and neutrophil extracellular traps (NET) formation.^38^ Our results argue against the role of neutrophils in a direct tissue injury, but they do not exclude neutrophils’ involvement in intravascular damage and cytokine storm driving.

Noteworthy, the boy who was excluded from the analysis due to the unclear character of his disease at the time of surgery did not have neutrophils in the appendix either. In addition to the presence of conjunctivitis, lymphopenia and substantially increased inflammatory markers, this finding might have facilitated earlier diagnosis of MIS-C in his case.

### 4.3. Lymphocyte subsets’ shifts in the appendices of MIS-C patients

A few case reports described lymphocytic infiltration of the intestine during MIS-C.^19,21^ The appendix is naturally rich in lymphocytes, but we found that appendiceal lymphocyte subpopulations differed in MIS-C compared to AA specimens. The proportion of B cells (CD20^+^) to T cells (CD5^+^) was higher in MIS-C appendices, which could be due to either B cell accumulation or T cell depletion (or both). Moreover, we observed the profound disproportion between CD4^+^ and CD8^+^ T cells, with the latter virtually absent in the appendices of MIS-C patients (median 5%). These findings closely resemble lymphocyte subpopulations’ shifts in the peripheral blood of children with MIS-C. Lymphopenia is a characteristic feature of MIS-C, with lymphocytes T more depleted than lymphocytes B, and CD8^+^ T cells being the most severely affected.^37,39,40^ Interestingly, decreased lymphocyte counts correlate with the disease severity.^37,39,40^ Deep immune serum profiling revealed that MIS-C is associated with an induction of cytokines and chemokines responsible for T cells’ recruitment from the circulation,^16,41^ which led to the hypothesis that lymphopenia of MIS-C is a result of lymphocytes’ shift from the blood to the tissues. Our finding of severely depleted CD8^+^ T cells in the appendices of MIS-C patients, parallel to decreased CD8^+^ T cells in the blood – as described in the literature, does not support this theory. However, to reliably assess lymphocyte migration to the peripheral tissues, specimens from other organs involved in MIS-C should be closely examined.

### 4.4. Absent SARS-CoV-2 antigens in the appendices of MIS-C patients

We found no SARS-CoV-2 nucleocapsid antigens in the appendices of children with MIS-C, which is contrary to the current hypotheses explaining MIS-C pathogenesis. It has been revealed that the SARS-CoV-2 spike protein includes superantigen motif, which could be responsible for uncontrolled immune activation and cytokine storm.^42–45^ However, it remains unclear why MIS-C develops with such a delay since the SARS-CoV-2 infection. Recently discovered high SARS-CoV-2 antigenemia in children with MIS-C, together with biomarkers of lost intestinal wall integrity, suggest that the intestine is a reservoir of SARS-CoV-2 antigens which leak into the circulation.^14^ Duarte et al. confirmed the presence of SARS-CoV-2 (by immunochemistry, PCR, and electron microscopy) in the intestine of a child who died from MIS-C.^19^ The same group found SARS-CoV-2 by PCR and immunohistochemistry in the appendix of a patient who underwent appendectomy during SARS-CoV-2 infection and suspected MIS-C.^20^ However, the latter patient did not manifest mucocutaneous or cardiovascular signs and recovered without immunomodulatory treatment, which questions the diagnosis of MIS-C. Interestingly, in another study, including nine children with AA and positive nasopharyngeal swab for SARS-CoV-2, the virus was absent in all specimens.^46^ Consistent with our results, Sahn et al. found no viral particles in the ileocolonic specimen from the child operated during MIS-C.^21^ To our knowledge, there are no more reports concerning the actual presence of SARS-CoV-2 antigens in the intestine of children with MIS-C, and our sample of 21 specimens revealing no SARS-CoV-2 nucleocapsid antigen in the appendiceal tissue is the largest described so far.

Another hypothesis involves T cell exhaustion due to persistent viral exposure.^18^ Thus, we investigated appendiceal tissue for PD-1, a marker of lymphocyte T exhaustion.^18,26^ We found no difference in PD-1 expression in the appendices from MIS-C and AA patients, and no PD-1^+^ lymphocytes were noticed outside germinal centers. It is consistent with the absence of SARS-CoV-2 antigens in the appendiceal tissue.

These findings do not reject the pathophysiological hypotheses listed above, but they argue against the appendix involvement in the viral persistence and subsequent immune dysregulation leading to MIS-C.

### 4.5. Limitations

Our study has several limitations. Firstly, CD5 is an imperfect marker of T lymphocytes, as it can also be present on immature B lymphocytes, and these are particularly common in the appendiceal wall.^47^ However, even if some of the CD5^+^ cells detected in our study were B cells instead of T cells, this would only additionally increase the ratio of B cells to T cells, which we found to be elevated in MIS-C. Secondly, the appendix is naturally rich in lymphatic tissue and thus not representative of other tissues. Therefore, we could not draw any conclusions about the inflammatory infiltration or hypothesized migration of lymphocytes to other organs typically involved in MIS-C (e.g., skin, and mucous membranes or myocardium) based on our results. Thirdly, the immunohistochemical finding of absent SARS-CoV-2 in the appendices of our MIS-C patients was not confirmed with other techniques, e.g., electron microscopy or PCR.

### 4.6. Conclusions

Despite frequent involvement of the gastrointestinal tract and clinical presentation of ‘acute abdomen’, the appendices of patients with MIS-C lack edema and neutrophilic infiltration of the muscularis. Lymphocyte subpopulations within the appendiceal wall mirror lymphocyte subpopulation shifts found in the blood during MIS-C. SARS-CoV-2 antigens are absent in the appendices of children with MIS-C. These findings argue against the central role of SARS-CoV-2 persistence in the gut and concomitant lymphocyte exhaustion as the major triggers of MIS-C.

## Data Availability

All data produced in the present study are available upon reasonable request to the authors.

## Abbreviations

AA: (acute appendicitis)
BMI: (body mass index)
BNP: (B-type natriuretic peptide)
COVID-19: (coronavirus disease 2019)
H&E: (hematoxylin and eosin)
HR: (heart rate)
INR: (international normalized ratio)
IVIG: (intravenous immunoglobulin)
LVEF: (left ventricular ejection fraction)
MIS-C: (multisystem inflammatory syndrome in children)
MOIS-CoR: (the MultiOrgan Inflammatory Syndromes Covid Related)
NT-proBNP: (N-terminal prohormone of B-type natriuretic peptide)
PCR: (polymerase chain reaction)
PCT: (procalcitonin)
PD-1: (programmed cell death 1 protein)
PICU: (pediatric intensive care unit)
SARS-CoV-2: (severe acute respiratory syndrome coronavirus 2)
sBP: (systolic blood pressure)
Tfh: (T follicular helper cells)
WBC: (white blood cells)
WHO: (World Health Organization)

